# Effectiveness of a bedaquiline, linezolid, clofazimine “core” for multidrug-resistant tuberculosis

**DOI:** 10.1101/2024.01.18.24301453

**Authors:** Chengbo Zeng, Miguel A. Hernán, Letizia Trevisi, Sara Sauer, Carole D. Mitnick, Catherine Hewison, Mathieu Bastard, Palwasha Khan, Kwonjune J. Seung, Michael L. Rich, Stephanie Law, Marina Kikvidze, Ohanna Kirakosyan, Alexey Miankou, Phone Thit, Shahid Mamsa, Aleeza Janmohamed, Nara Melikyan, Saman Ahmed, Dante Vargas, Amsalu Bekele Binegdie, Kulbaram Temirova, Lawrence Oyewusi, Kerline Philippe, Stalz C. Vilbrun, Uzma Khan, Helena Huerga, Molly F. Franke, endTB Observational Study Team

## Abstract

**Rationale:** Treatment outcomes may be compromised among patients with multidrug- or rifampicin-resistant tuberculosis with additional fluoroquinolone resistance. Evidence is needed to inform optimal treatment for these patients.

**Objectives:** We compared the effectiveness of longer individualized regimens comprised of bedaquiline for 5 to 8 months, linezolid, and clofazimine to those reinforced with at least 1 third-tier drug and/or longer duration of bedaquiline.

**Methods:** We emulated a target trial to compare the effectiveness of initiating and remaining on the core regimen to one of five regimens reinforced with (1) bedaquiline for ≥9 months, (2) bedaquiline for ≥9 months and delamanid, (3) imipenem, (4) a second-line injectable, or (5) delamanid and imipenem. We included patients in whom a fluoroquinolone was unlikely to be effective based on drug susceptibility testing and/or prior exposure. Our analysis consisted of cloning, censoring, and inverse-probability weighting to estimate the probability of successful treatment.

**Measurements and Main Results:** Adjusted probabilities of successful treatment were high across regimens, ranging from 0.75 (95%CI:0.61, 0.89) to 0.84 (95%CI:0.76, 0.91). We found no substantial evidence that any of the reinforced regimens improved effectiveness of the core regimen, with ratios of treatment success ranging from 1.01 for regimens reinforced with bedaquiline ≥9 months (95%CI:0.79, 1.28) and bedaquiline ≥9 months plus delamanid (95%CI:0.81, 1.31) to 1.11 for regimens reinforced by a second-line injectable (95%CI:0.92, 1.39) and delamanid and imipenem (95%CI:0.90, 1.41).

**Conclusions:** High treatment success underscores the effectiveness of regimens comprised of bedaquiline, linezolid, and clofazimine, highlighting the need for expanded access to these drugs.

## Introduction

The use of new and repurposed drugs, such as bedaquiline (Bdq), delamanid (Dlm), linezolid (Lzd), pretomanid, and clofazimine (Cfz), has drastically improved the effectiveness of treatment for multidrug- or rifampicin-resistant tuberculosis (MDR/RR-TB).^1–5^ For the first time in history, the recommended treatment duration for MDR/RR-TB is as short as six-to-nine months for the majority of patients.^6–10^ However, longer 18-20 month regimens are still recommended in cases where shorter regimens cannot be used (i.e., due to confirmed or suspected drug resistance and/or unavailability of drugs in the shorter regimens).^6^ Key research priorities highlighted by the World Health Organization (WHO) with regard to longer MDR/RR-TB regimens are studies on the optimal number and combination of drugs for patients previously treated for RR/MDR-TB; the approach to regimen design; and the optimal duration of Bdq.^6^

Patients previously treated for MDR/RR-TB, are at increased risk for unfavorable treatment outcomes, in part due to a higher risk of resistance to fluoroquinolone drugs, a cornerstone of longer individualized treatments for MDR/RR-TB.^1,11^ In this paper, we sought to address knowledge gaps with regard to the optimal treatment for patients requiring treatment for MDR/RR-TB, in whom a fluoroquinolone (FQ) is unlikely to be effective. Specifically, we emulated a target trial to compare the effectiveness of a “core” regimen comprised of Bdq for 5 to 8 months, Lzd, and Cfz with regimens that were reinforced with at least one third tier (i.e., Group C) drug and/or a longer duration of Bdq.

## Methods

### Data resources and study population

The prospective endTB observational cohort (NCT03259269) aimed to generate evidence on the safety and effectiveness of Bdq or Dlm when used as part of a longer multidrug regimen for RR/MDR-TB.^12^ The cohort includes 2788 patients from 17 countries who initiated a Bdq- or Dlm-containing regimen between April 2015 and September 2018 and consent to be enrolled. Each participant was followed according to local program norms. Data were collected using standardized forms and entered into an electronic medical record. For this analysis, we excluded participants from the Democratic People’s Republic of Korea (DPRK) due to differences in diagnosis and treatment compared with the rest of the cohort.

### Specification of the target trial

The (hypothetical) pragmatic trial would enroll participants within a week of MDR/RR-TB treatment initiation in whom a FQ is unlikely to be effective and in whom the following drugs are likely to be effective: Bdq, Lzd, Cfz, Dlm, Imipenem (Imp), and at least one second-line injectable (SLI) drug (i.e., Capreomycin [Cm], Kanamycin [Km], and Amikacin [Am]). The likely-effectiveness of a drug in an individual is based on drug susceptibility testing (DST) or prior history of these drugs (if no DST was available).

Two weeks after enrollment, each eligible individual would be randomly assigned to one of six treatment strategies: the core regimen (Bdq for 5 to 8 months, Lzd, Cfz) or one of five reinforced regimens: (1) Bdq (≥ 9 months)-Lzd-Cfz; (2) Bdq (≥ 9 months)-Lzd-Cfz-Dlm; (3) Bdq (≥ 5 months)-Lzd-Cfz-Imp; (4) Bdq (≥ 5 months)-Lzd-Cfz-SLI; (5) Bdq (≥ 9 months)-Lzd-Cfz-Dlm-Imp (Online Data Supplement Table E1). Lzd is initiated at a dose of 600mg daily but can be reduced to other doses if clinically indicated. Regimens are intended to last 18 to 20 months, but clinicians determine the total duration of treatment and of each individual drug, except Bdq, which is protocolized into one of three durations: ≥ 5 months, 5-8 months, ≥ 9 months. Clinician-directed Bdq interruptions of less than or equal to 14 days are allowable for any reason (e.g., toxicity, drug stock-out). Bdq suspensions of longer than 14 days and additions of any drug for longer than 14 days are not permitted unless in response to an adverse event (AE) or acquired-resistance to a drug in the assigned treatment strategy. Bdq can be reinitiated after the stoppage indicated by the assigned strategy, if clinically indicated, and drugs that are unlikely effective can be included in the regimen.

The outcome of interest is treatment success, defined as cure or treatment completion at the end of treatment. Death, treatment failure, and loss to follow-up are considered unsuccessful end-of-treatment (EOT) outcomes. All EOT outcomes are calculated based on the WHO guidance and identify the first point at which failure occurs.^13,14^ For each individual, follow-up would start at assignment to a regimen (time 0) and continue each week until the end of treatment. The causal contrasts of interest are the intention-to-treat effect and the per-protocol effect.

### Statistical analysis of the target trial

In the intention-to-treat analysis, the probabilities of treatment success in each group can be estimated nonparametrically or via a parametric logistic regression model for the weekly probability of treatment success (the model can include baseline covariates if their distribution differs between groups). The predicted probabilities of treatment success are then compared via success ratios and differences. The 95% confidence intervals are computed using the bootstrapping method with 500 samples.

The per-protocol analysis is identical except that individuals are censored if/when their treatments deviated from their assigned strategy for any reasons other than adverse effects or acquired-resistance to a drug in the assigned strategy. Specifically, individuals are censored if they have any new likely-effective drugs added to their assigned strategy or if they do not follow the assigned duration of Bdq. To adjust for the potential selection bias introduced by censoring, we can incorporate inverse-probability (IP) weights.

### Target trial emulation

We emulated the target trial using the endTB prospective observational dataset (Online Data Supplement Table E1 and Figure E1).^15,16^ Inclusion criteria were the same, except that eligible individuals were those in whom *at least* Bdq, Lzd, and Cfz were likely to be effective (i.e., likely-effectiveness to a SLI, Dlm and/or Imp was not required for inclusion). Because individuals can have data compatible with more than one treatment strategy at time 0, we made one modification to the per-protocol analysis^15,17^: we cloned individuals in the dataset and assigned each clone to each of the Bdq durations (i.e., Bdq ≥ 5 months, Bdq 5 to 8 months, or Bdq ≥ 9 months) that were compatible with their observed data at time 0. Online supplement Figure E2 shows an overview of the cloning and censoring steps. We fit an IP-weighted logistic regression model for the probability of treatment success among uncensored clones that included the following baseline covariates^18^: treated in Georgia (yes/no), year of enrollment (continuous), low BMI (yes/no), sputum smear (positive/negative), sputum culture (positive/negative), and receiving cycloserine (Cs), although it was unlikely to be effective in the individual (yes/no).

We estimated the denominator of the stabilized IP weights using separate logistic models for the weekly probability of “not adding any new likely-effective drugs to the baseline regimens” and of “remaining on Bdq” conditional on baseline and time-varying covariates. Time-varying confounders were sputum smear (positive/negative) and receipt of Cs which was unlikely to be effective in the individual (yes/no). Time was modelled with linear and quadratic terms for the week of follow-up. We estimated the numerator of the stabilized IP weights using analogous models without time-varying covariates. Online supplement Table E2 provides the calculation of IP weights for each treatment strategy. IP weights were not truncated.

### Research ethics

The endTB observational study protocol was approved by all study countries and central ethics review committees for each consortium partner (Partners In Health, Doctors Without Borders, Epicentre, and Interactive Research and Development). Participants provided written informed consent for inclusion in the observational cohort.

## Results

Of 446 eligible patients across 13 countries, 443 had a known EOT outcome and were included in our analysis (Online Data Supplement Figure E1). One-third (149/443) of the participants were female, and the median age was 35 (IQR: 28, 45) (Table 1). The prevalence of comorbidities was: 4% HIV infection, 12% diabetes mellitus or glucose intolerance, 6% hepatitis B infection, and 17% hepatitis C infection. Also, 96% had resistance to a FQ, 69% had bilateral disease, and 75% had cavitary disease.

**Table 1.** Baseline characteristics of 443 individuals receiving treatment for MDR/RR-TB who were previously treated with second-line tuberculosis drugs, 2015-2018.

| Characteristics | n (%)<sup>*</sup> |
| --- | --- |
| Demographic characteristics |  |
| Age at treatment initiation (year, median, IQR, range) | 35 (28, 45; 14, 71) |
| Female | 149 (34) |
| Marital status, married or living together | 211 (48) |
| Substance and drug use |  |
| Alcohol use | 55 (12) |
| Tobacco use (N=441) | 142 (32) |
| Injection drug use (N=441) | 6 (1) |
| Non injection drug use (N=439) | 14 (3) |
| Comorbidities |  |
| Anemia (N=439) | 181 (41) |
| HIV infection | 19 (4) |
| Diabetes mellitus or glucose intolerance (N=442) | 53 (12) |
| Hepatitis B virus infection (N=442) | 28 (6) |
| Hepatitis C virus infection | 77 (17) |
| At least one comorbidity other than those above | 48 (11) |
| TB-related characteristics |  |
| Prior TB treatment with second-line drugs | 439 (99) |
| Bilateral disease (N=427) | 296 (69) |
| Cavitary disease (N=417) | 313 (75) |
| Positive culture (N=412) | 276 (67) |
| Positive smear (N=423) | 250 (59) |
| Resistance profile |  |
| MDR/RR-TB without any testing to FQ | 33 (7) |
| MDR/RR-TB with resistance to any FQ | 410 (96) |
| Numbers of not likely effective drugs included in baseline regimen (median, IQR, range) | 1 (0, 2; 0, 5) |
| Impaired functional status (limited self-care or completely disabled) (N=377) | 192 (51) |
| Body mass index <18.5 (N=442) | 154 (35) |

Abbreviations: MDR/RR-TB: Multidrug- or rifampicin- resistant tuberculosis. FQ:
Fluoroquinolone. \*: Unless otherwise noted. IQR: Interquartile range.

Of the 443 individuals, 23% (n=100) initiated a regimen containing Bdq, Lzd, and Cfz, without additional likely effective drugs. The remaining individuals received one or more likely effective third-tier drugs, including Dlm (18%, n=81), Imp (17%, n=74), SLI (23%, n=103), and Dlm and Imp together (19%, n=85). All but 1 individuals (0.2%) initiated Lzd at a dose of 600 mg daily. The proportion who received Cs that was unlikely effective for them was 65% (65/100) among participants who initiated Bdq-Lzd-Cfz and 67% (69/103) among those initiating a reinforced regimen containing injectable. In other regimens, the receipt of Cs ranged from 6 (Bdq-Lzd-Cfz-Dlm-Imp: 5/85) to 36% (Bdq-Lzd-Cfz-Imp: 27/74; Online Data Supplement Table E3).

The probability of treatment success ranged from 0.75 (95%CI: 0.61, 0.89) for Bdq (5 to 8 months)-Lzd-Cfz to 0.84 (95%CI: 0.76, 0.91) for Bdq (≥5 months)-Lzd-Cfz-SLI (Table 2). Compared with Bdq (5 to 8 months)-Lzd-Cfz, the treatment success ratios ranged from 1.01 for regimens reinforced with bedaquiline ≥9 months (95%CI:0.79, 1.28) and bedaquiline ≥9 months plus delamanid (95%CI:0.81, 1.31) to 1.11 for regimens reinforced by a second-line injectable (95%CI:0.92, 1.39) and by delamanid and imipenem (95%CI:0.90, 1.41) (Table 2). The risk difference ranged from 0.01 for regimens reinforced with bedaquiline ≥9 months (95%CI:−0.18, 0.17) and bedaquiline ≥9 months plus delamanid (95%CI:−0.16, 0.19) to 0.08 for regimens reinforced by a second-line injectable (95%CI:−0.08, 0.23) and by delamanid and imipenem (95%CI:−0.08, 0.26) (Figure 1). The mean of stabilized IP weights was 1.00 (standard deviation: 0.13; range: 0.71, 4.20).

**Figure 1.**
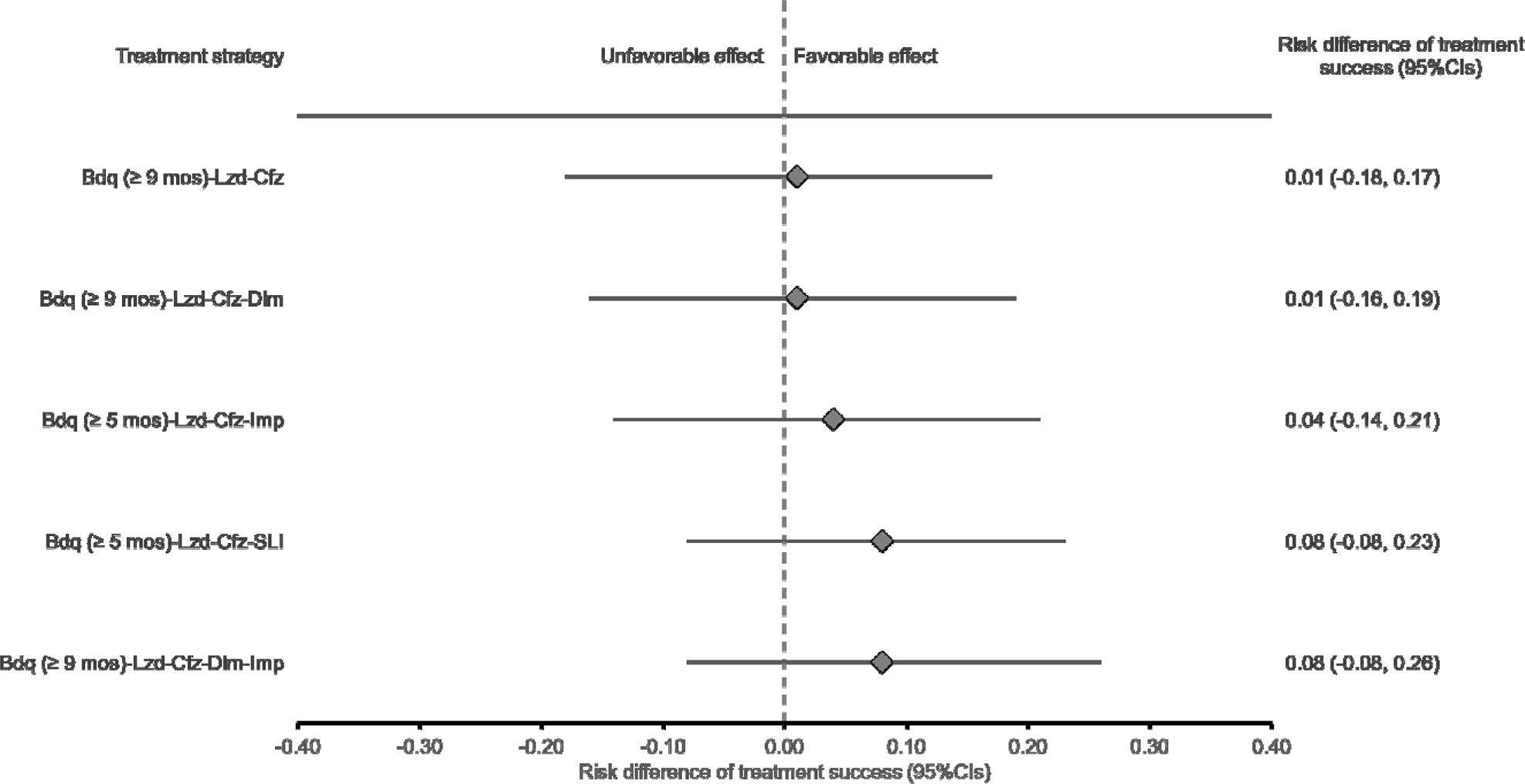
Adjusted risk differences for treatment success of reinforced regimens compared to the Bdq (5 to 8 months)-Lzd-Cfz core regimen. Abbreviations: Bdq: Bedaquiline. Lzd: Linezolid. Cfz: Clofazimine. Dlm: Delamanid. Imp: Imipenem. SLI: Second-line injectable. CI: Confidence interval.

**Table 2.** Adjusted probabilities and ratios of MDR/RR-TB end-of-treatment success among individuals previously treated with second-line tuberculosis drugs.

| Treatment strategy | Adjusted probability of<br>treatment success<br>(95%CI) | Adjusted ratio of<br>treatment success<br>(95%CI) |
| --- | --- | --- |
| Bdq-(5 to 8 mos)-Lzd-Cfz | 0.75 (0.61, 0.89) | Ref. |
| Bdq-(≥ 9 mos)-Lzd-Cfz | 0.77 (0.64, 0.87) | 1.01 (0.79, 1.28) |
| Bdq-(≥ 9 mos)-Lzd-Cfz-Dlm | 0.76 (0.65, 0.87) | 1.01 (0.81, 1.31) |
| Bdq-(≥ 5 mos)-Lzd-Cfz-Imp | 0.80 (0.68, 0.90) | 1.06 (0.83, 1.34) |
| Bdq-(≥ 5 mos)-Lzd-Cfz-SLI | 0.84 (0.76, 0.91) | 1.11 (0.92, 1.39) |
| Bdq-(≥ 9 mos)-Lzd-Cfz-Dlm-Imp | 0.83 (0.74, 0.91) | 1.11 (0.90, 1.41) |

Abbreviations: MDR/RR-TB: Multidrug- or rifampicin- resistant tuberculosis. Bdq: Bedaquiline.
Lzd: Linezolid. Cfz: Clofazimine. Dlm: Delamanid. Imp: Imipenem. SLI: Second-line injectable.
mos: Months. CI: Confidence interval.
Adjusted for year of enrollment (i.e., 2015, 2016, 2017, 2018), treated in Georgia (yes/no), low BMI (yes/no), sputum smear (negative/positive), time-varying sputum smear (negative/positive), sputum culture (negative/positive), received cycloserine that was unlikely to be effective (yes/no), time-varying receipt of cycloserine (yes/no), and missing indicator of sputum culture.

## Discussion

In a cohort of individuals with MDR/RR-TB in whom a FQ was unlikely to be effective, longer regimens containing at least Bdq for five months, Lzd, and Cfz yielded high proportions of treatment success that align with those from randomized trials and observational cohorts of patients treated with longer regimens comprised of new and repurposed drugs.^1–5,8^ The regimens reinforced with at least one Group C drug and/or a longer duration of Bdq did not substantially improve the frequency of treatment success compared with the Bdq (5 to 8 months)-Lzd-Cfz core regimen. This finding underscores that Bdq (5 to 8 months)-Lzd-Cfz was a potent regimen for treating individuals with MDR/RR-TB in whom a FQ was unlikely to be effective. Lzd was initiated at a dose of 600 mg daily, which likely contributed to the effectiveness of the core regimen.^7^ Prior studies have highlighted the challenges of evaluating the effectiveness of adding a single drug to a highly effective regimen.^19,20^

Use of a target trial framework to address this research question facilitated inference and interpretation in several regards. First, it enabled clear articulation of the research question and the treatment strategies to be compared. Second, it informed the design of an analysis that accounted for two important sources of potential bias: (1) regimens were assigned and changed based on clinical judgment, not at random, which can lead to important differences among the individuals who receive each regimen (i.e., confounding); (2) individuals who live longer can be treated longer (i.e., immortal person-time bias).^17,21^ Although both biases are important considerations in observational comparative effectiveness analyses of treatment duration, methods to appropriately account for them have been applied infrequently in TB cohorts.^4,21–23^

The 2022 WHO guidance recommends that longer regimens for MDR/RR-TB prioritize drugs from Groups A (Bdq, Lzd, FQ) and B (Cfz and Cs).^6^ Under this guidance, the recommended longer regimen for someone with FQ-resistant TB consists of at least the four-drug regimen Bdq-Lzd-Cfz-Cs. We were unable to study this regimen because Cs rarely met the definition of a likely-effective drug in this cohort of individuals previously exposed to second-line drugs. Therefore, the core combination of Bdq-Lzd-Cfz plus Cs, where all four drugs were likely effective, could not be evaluated in this study.

Although we observed a moderately higher frequency of treatment success with three regimens, one reinforced with Imp, one reinforced with a SLI, and one reinforced with Dlm, Imp and longer Bdq duration, 95% confidence intervals around these estimates were wide. These modest increases in effectiveness may be due to true differences, chance, or residual confounding.^24^ The relatively small sample size for this analysis forced us to balance confounder adjustment and model stability, and therefore we focused on the strongest and most likely confounders. Therefore, residual confounding is a possibility. We used the missing indicator method to account for missing data on confounders, an approach that retains the original analytic sample size but cannot completely adjust for confounding.^25^

One example of residual confounding is that regimen composition is often clustered by study sites based on the national TB program guidelines. For example, the regimen reinforced with an injectable was disproportionately used in Georgia; we adjusted for this site, but the sample size precluded us from adjusting for all 13 sites included in this analysis. Addressing this question within a larger longitudinal cohort could facilitate more conclusive findings in several ways. First, it would increase statistical power and reduce sampling variability. Second, it would afford greater flexibility to adjust for all potential confounders. Third, it would facilitate the potential for comparisons in key subgroups, such as individuals living with HIV.

In conclusion, among individuals with MDR/RR-TB in whom a FQ is unlikely to be effective due to resistance or prior exposure to the second-line treatment, we found high rates of treatment success but no conclusive evidence that reinforcing a core regimen of Bdq (5 to 8 months)-Lzd-Cfz with additional Group C drugs and/or a longer duration of Bdq improved the treatment effectiveness. The high treatment success underscores the need for expanded access to these drugs and supports the effectiveness of current WHO drug hierarchy for the design of longer regimens. Future evaluations could examine alternative strategies for optimizing this and other core regimens and informing management strategies for suspension and discontinuation of core drugs. Analyses will be improved through larger longitudinal cohorts, which will enhance precision, enable more complete control of potential biases and permit examination of subgroup heterogeneity.

## Data Availability

All data produced in the present study are available upon reasonable request to the authors.

## Author contributions

Study design: KJS, HH, UK, MLR, CDM, CH, MB, PYK, MFF

Analysis design: MAH, MFF, LT, CZ

Study implementation: MK, OK, AM, PT, SM, AJ, NM, SA, DV, ABB, KT, LO, KP, SCV

Data management: CZ, LT

Statistical analysis: CZ, SS, MFF

Data interpretation: CZ, MFF, LT, MAH, CDM, MLR, KJS, UK, PYK, CH, SL, SS

First draft: CZ, MFF

Critical review of manuscript: all authors

## Conflicts of Interest and Source of Funding

Bedaquiline donations made from Janssen to the Global Drug Facility were used for patients in the endTB observational study. Donations of delamanid from Otsuka were used for initial patients enrolled in the endTB observational study. The donors did not have any role in the study design, data analyses, data interpretation or manuscript writing. All authors report no additional potential conflicts of interest. The endTB observational study was funded by UNITAID. MFF, CZ, MAH, LT, MLR, KJS and CDM were supported by the National Institute of Allergy and Infectious Diseases of the National Institutes of Health under Award Number R01AI46095. The funders had no role in the conceptualization, analysis or presentation of findings of this study.

## Acknowledgements

The authors thank the participants for their participation in the endTB Observational Study and the clinicians and program staff for their support on recruitment, operation, and management. The authors also thank endTB staff at Partners In Health, Doctors Without Borders, Epicentre, and Interactive Research and Development.

## Online Supplement Materials

**Table E1.** Protocol of comparative effectiveness analysis of a regimen comprised of Bdq-(5 to 8 months)-Lzd-Cfz, among individuals in whom a FQ was unlikely to be effective.

| Component | Hypothetical clinical trial | Emulated trial using endTB observational cohort |
| --- | --- | --- |
| **Eligibility criteria** | - Confirmed MDR/RR-TB<br>- Started Bdq within a week of treatment initiation<br>- Fluoroquinolone unlikely to be effective<br>- Likely-effectiveness of Bdq, Lzd, Cfz, Dlm, Imp, and at least one SLI drug (Cm, Km, and Am) | Same except that individuals had likely effectiveness to at least Bdq, Lzd, and Cfz. They could also have likely-effectiveness to at least one SLI or Dlm and/or Imp |
| **Treatment strategies** | 0. Bdq (5 to 8 months)-Lzd-Cfz<br>1. Bdq ($\geq 9$ months)-Lzd-Cfz<br>2. Bdq ($\geq 9$ months)-Lzd-Cfz-Dlm<br>3. Bdq ($\geq 5$ months)-Lzd-Cfz-Imp<br>4. Bdq ($\geq 5$ months)-Lzd-Cfz-SLI<br>5. Bdq ($\geq 9$ months)-Lzd-Cfz-Dlm-Imp<br>- Regimens are intended to last 18 to 20 | Same |
|  | months, but clinicians determine the total duration of treatment and of each individual drug, except Bdq, which is protocolized into one of three durations: $\geq 5$ months, 5-8 months, and $\geq 9$ months.<br>- Clinician-directed drug interruptions of less than or equal to 14 days were allowable for any reason (e.g., toxicity, drug stock-out).<br>- Drug additions or suspensions of longer than 14 days are not permitted except in response to an AE or acquired resistance to a drug in the assigned treatment strategy.<br>- Bdq can be reinitiated after the stoppage indicated by the assigned strategy, if clinically indicated.<br>- Drugs that are unlikely to be effective can be included in the regimen. |  |
| **Treatment assignment** | Individuals are randomly assigned to one of the six treatment strategies on day 14 of treatment and are aware of their assignment. | Individuals are assigned to the treatment strategies that their observed data are compatible with at day 14 of treatment. |
| **Follow-up period** | Follow-up starts at assignment and continues | Same |
|  | until the occurrence of an end-of-treatment outcome. |  |
| **Outcome** | Treatment success (i.e., cure or treatment completion). Individuals who die, become lost to follow-up, or in whom treatment fails would be considered to have unsuccessful treatment outcomes. | Same |
| **Causal contrast of interest** | Intention-to-treat and per-protocol effects | Observational analogue of the per-protocol effect |
| **Analysis plan** | Per-protocol effect: Individuals are censored if their treatment deviates from their assigned strategy, and potential selection bias introduced by censoring is adjusted for via IP weighting. Estimates are adjusted for baseline covariates that differ across groups. Probabilities of treatment success are compared via ratios and differences. | Same except that person clones are used. |

Abbreviations: FQ: Fluoroquinolone. MDR/RR-TB: Multidrug- or rifampicin-resistant tuberculosis. AE: Adverse event. Bdq:
Bedaquiline. Lzd: Linezolid. Cfz: Clofazimine. Dlm: Delamanid. Imp: Imipenem. SLI: Second-line injectable. Cm: Capreomycin. Km:
Kanamycin. Am: Amikacin. IP: Inverse-probability.

**Table E2.** Probabilities of being uncensored for each Bdq duration strategy and follow-up period.

| Bdq duration strategy | Probabilities of being uncensored |  |  |  |
| --- | --- | --- | --- | --- |
|  | 0 < t < 5-month | 5-month ≤ t ≤ 8-month | 9-month | t > 9-month |
| **At least 5 months** |  |  |  |  |
| Bdq, Lzd, Cfz, Imp | 1*Prob. no added drugs | 1*Prob. no added drugs | 1*Prob. no added drugs | 1*Prob. no added drugs |
| Bdq, Lzd, Cfz, SLI | 1*Prob. no added drugs | 1*Prob. no added drugs | 1*Prob. no added drugs | 1*Prob. no added drugs |
| **5 to 8 months** |  |  |  |  |
| Bdq, Lzd, Cfz | 1*Prob. no added drugs | 1*Prob. no added drugs | (1-Prob. on Bdq for those who had never stopped Bdq before<sup>□</sup>)*<br>Prob. no added drugs | 1*Prob. no added drugs |
| **At least 9 months** |  |  |  |  |
| Bdq, Lzd, Cfz | 1*Prob. no added drugs | Prob. on Bdq*Prob. no added drugs | 1*Prob. no added drugs | 1*Prob. no added drugs |
| Bdq, Lzd, Cfz, Dlm | 1*Prob. no added drugs | Prob. on Bdq*Prob. no added drugs | 1*Prob. no added drugs | 1*Prob. no added drugs |
| Bdq, Lzd, Cfz, Dlm, Imp | 1*Prob. no added drugs | Prob. on Bdq*Prob. no added drugs | 1*Prob. no added drugs | 1*Prob. no added drugs |

Abbreviations: Bdq: Bedaquiline. Lzd: Linezolid. Cfz: Clofazimine. Dlm: Delamanid. Imp: Imipenem. SLI: Second-line injectable. Prob:
Probability. t: Time. <sup>□</sup> : For other individuals, it is “1\*Prob. no added drugs” .

The probability of being uncensored is calculated for each week. At time 0, the probability of being uncensored (*p_0_*) is 1. Subsequently, for each week *i*, the overall probability of being uncensored (*P_i_*) is the cumulative product of the conditional probabilities of being uncensored in each week up to and including week *i*, i.e. *p_0_*,…, *p_i_* (Table E2). The formula for calculating *P_i_* is shown in Formula 1:

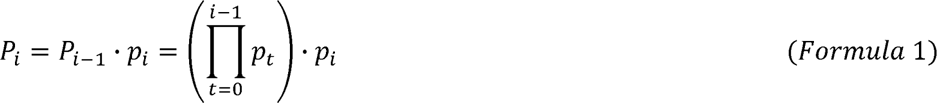

For each week, the weight (*W_i_*) is calculated as the inverse of the probability of being uncensored (*P_i_*) (Formula 2). The final weight at the end of treatment was retained for each individual and adjusted in the analysis.

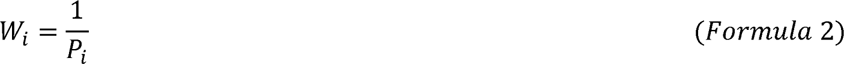

**Table E3.** Inclusion of cycloserine that was unlikely to be effective in regimens of 443 eligible individuals treated for MDR/RR-TB (day 14)

| Treatment strategy | No use of Cs | Use of Cs that was unlikely to be effective |
| --- | --- | --- |
| Bdq-Lzd-Cfz | 34 (34) | 65 (65) |
| Bdq-Lzd-Cfz-Dlm | 56 (69) | 25 (31) |
| Bdq-Lzd-Cfz-Imp | 47 (64) | 27 (36) |
| Bdq-Lzd-Cfz-SLI | 34 (33) | 69 (67) |
| Bdq-Lzd-Cfz-Dlm-Imp | 80 (94) | 5 (6) |

Abbreviations: Cs: Cycloserine. Bdq: Bedaquiline. Lzd: Linezolid. Cfz: Clofazimine. Dlm: Delamanid. Imp: Imipenem. SLI: Second-line injectable.
Dlm: Delamanid. Imp: Imipenem.

**Figure E1.**
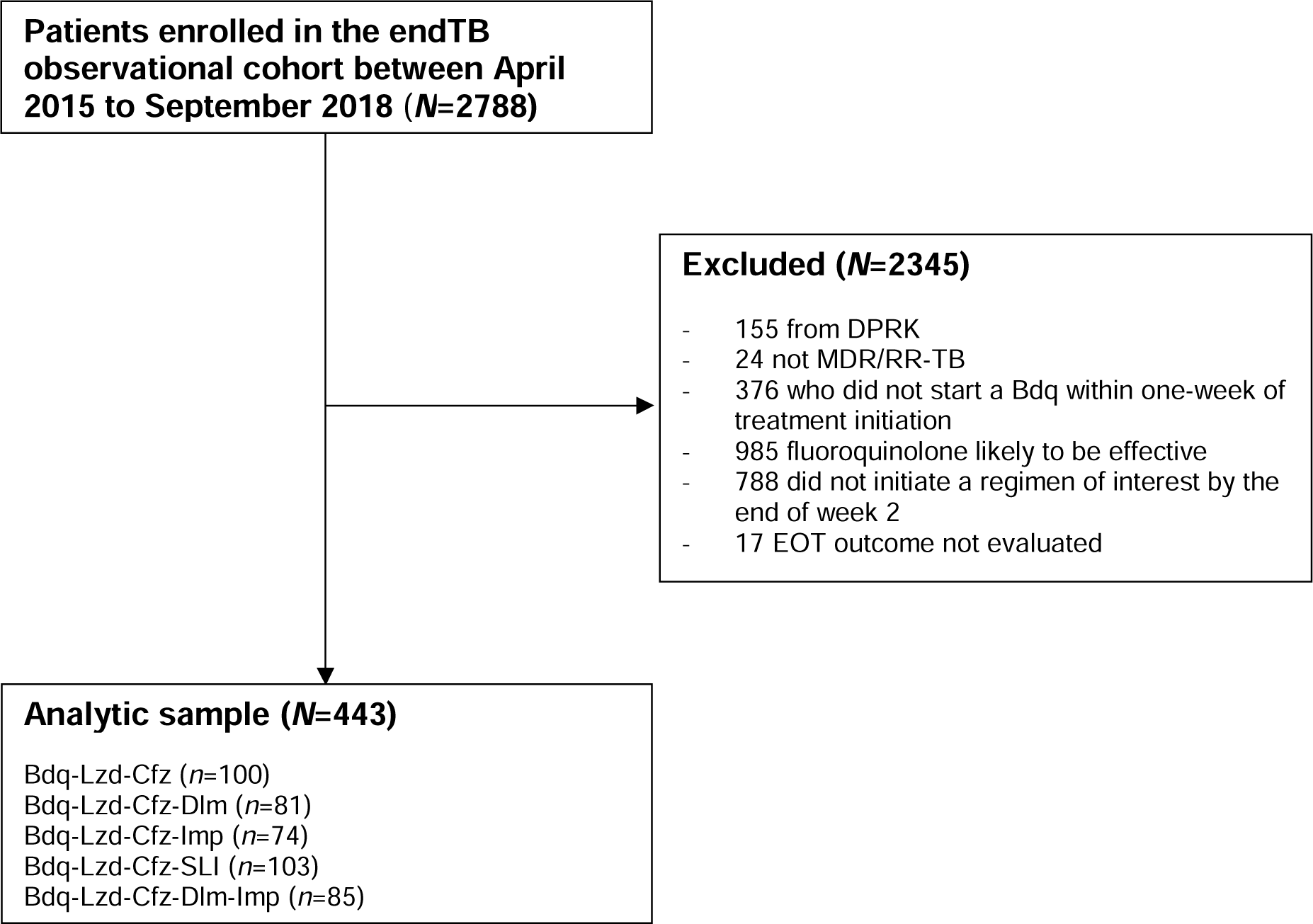
Inclusion flowchart of endTB Observational Study individuals included in the present analysis. Abbreviations: DPRK: Democratic People’s Republic of Korea. MDR/RR-TB: Multidrug- or rifampicin-resistant tuberculosis. EOT: End-of-treatment. Bdq: Bedaquiline. Lzd: Linezolid. Cfz: Clofazimine. Dlm: Delamanid. Imp: Imipenem. SLI: Second-line injectable.

**Figure E2.**
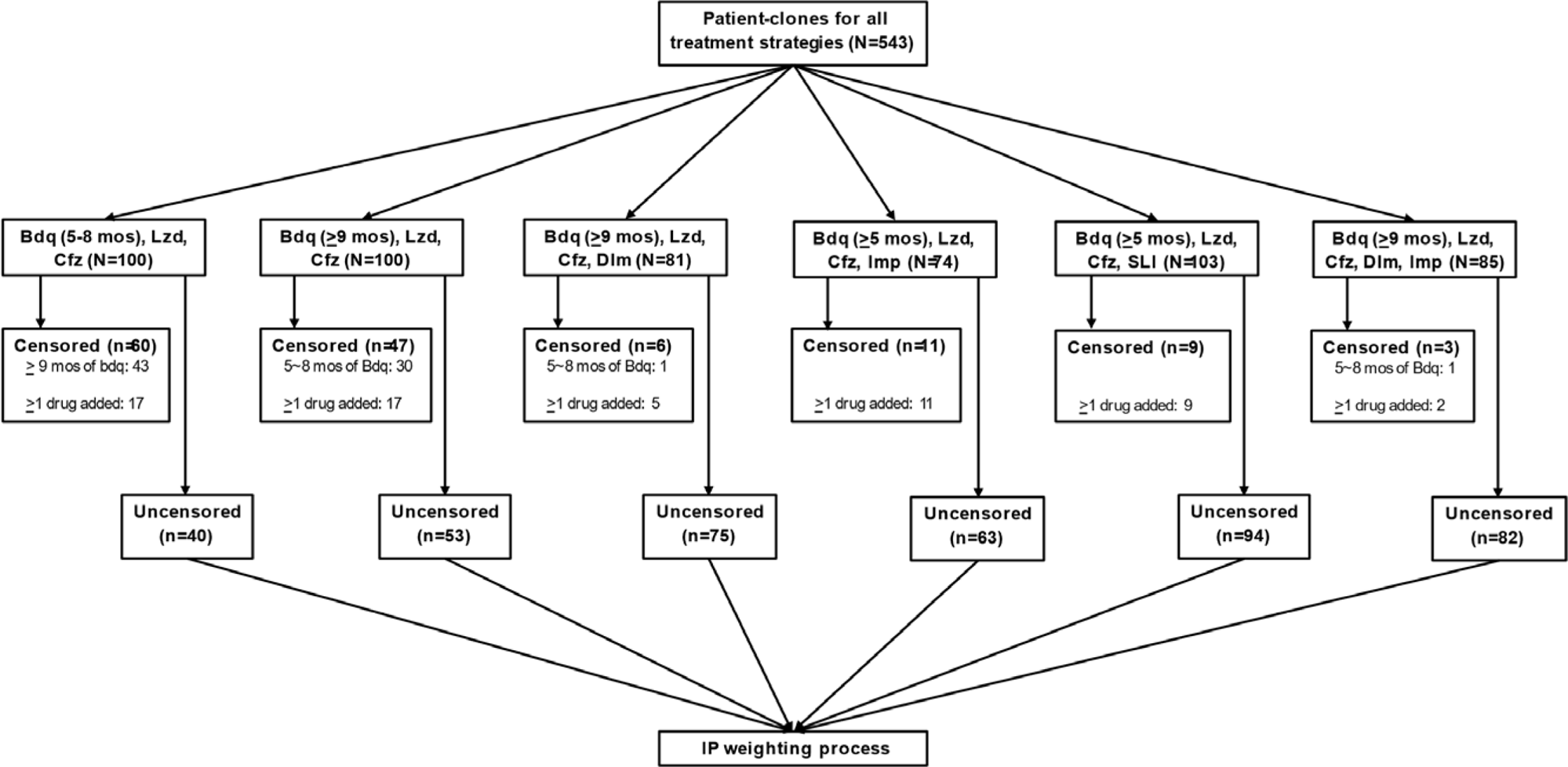
Cloning, censoring, and IP weighting processes for each treatment strategy. Abbreviations: mos: Months. SLI: Second-line injectable. IP: Inverse-probability. Bdq: Bedaquiline. Lzd: Linezolid. Cfz: Clofazimine. Dlm: Delamanid. Imp: Imipenem. Clones were censored if their treatment deviates from their assigned strategy; the uncensored clones were used to calculate the IP weights.

